# The Uptake of Critical Perspectives in the Field of Global Mental Health: A Critical Interpretive Synthesis

**DOI:** 10.1101/2024.01.10.24301044

**Authors:** Gojjam Limenih, Elysee Nouvet

## Abstract

An emancipatory movement is surging within the of global mental health (GMH) field, advancing the groundwork laid by scholars who champion a more critical, self-reflective, decolonized, and socially responsive discipline. As GMH enters its second decade, there have been numerous critiques of its origins and underlying paradigms, with increasing emphasis on critical approaches in recent publications. However, there is no comprehensive synthesis has yet been undertaken to understand how critical perspectives have been integrated into the GMH literature. To contribute to the ongoing discourse aimed at cultivating GMH as a critical and socially responsive discipline, this article employed the critical interpretive synthesis method. critical interpretive synthesis is designed to navigate the synthesis of extensive and diverse literature while actively engaging with the foundational assumptions that shape and inform the body of research via employing a critical lens to scrutinize the data. We conducted searches using PubMed, MEDLINE(OVID), PsycINFO, Scopus and EMBASE data bases; published between 2007 and February 2023. We included 58 articles that have embraced critical perspectives, whether explicitly or implicitly. Through this iterative process, five distinct themes or “four turns” emerge: (1) the inward turn, focusing on the “local” as a source of alterity, resistance, and critique; (2) turning the critical lens outward or political turn; (3) the push for a broader agenda; and (4) the reflexive turn. This article discusses the implications of four “turns” in how critical perspectives have been and are being used, in relation to the goal of developing as a socially responsive discipline.

## Introduction

Global mental health (GMH) is at a crucial moment of disciplinary development which is characterized by critical reflexivity about its foundational assumptions and calls for the field to become a more critical and “socially responsive” discipline (Bracken et al., 2016; Bemme & Kirmayer, 2020; Bemme, 2023). As the field enters its second decade of formal existence, the boundaries, and underlying assumptions within which it was built, as well as the type of knowledge it produces, have been increasingly expanded, examined, critiqued, and scrutinized (Bracken et al., 2016; Bemme & Kirmayer, 2020; Ecks, 2021; Kirmayer & Pederson, 2014; Mills, 2015; Summerfield, 2013). Several scholars have raised concerns regarding the dominance of a Eurocentric conceptualization of mental health and wellbeing, informed by ideas dominant in the “Western” world (Bracken et al., 2016; Ecks & Bassu, 2014; Ecks, 2021; Ingleby, 2014; Mills, 2014; Summerfield, 2006, 2008, 2013; Kirmayer & Pedersen, 2014), pointing to the limitations that this orientation imposes when attempting to understand the diversity of ways mental health, illness, and wellbeing are understood and enacted globally; but specifically in the Global South.

This shift has been coupled with growing awareness analysis of historical (colonial), political, social, economic, and pharmaceutical forces that extend beyond the individual and shape possibilities for people’s emotional wellbeing and health-care systems in the Global South (Burgess et al., 2020; Mills, 2015, 2022; Fernando, 2014, 2017; Kirmayer & Crafa, 2014; Kirmayer & Swartz, 2013; Tribe, 2014; White & Mills, 2017). This has led expanded understanding and shift the attention to social and cultural elements in addition to medical perspective. This shift has highlighted the importance of addressing political and structural inequalities (Applbaum, 2015; Farmer et al. 2006; Kirmayer & Pedersen, 2014; Lund et al., 2011; Marmot 2014; Tribe, 2014). Similarly, calls to address global inequities through the development of a more critical and socially responsive discipline, have also materialized (Bracken et al., 2016; Bemme & D’Souza, 2014; Bemme, 2019; Bemme & Kirmayer, 2020).

The necessity of such disciplinary development has involved scholars advocating an ‘emancipatory agenda’ (Bracken et al., 2016; Whitaker etal., 2021) and social reform in which the implicit power and influence of global pharmaceuticals is stressed (Applbaum, 2015) and the importance of fostering reflexive deliberation in practice is advocated (Gómez-Carrill et al., 2020) to develop a politically engaged and socially transformative discipline.

As noted by Bemme and Kirmayer (2020), there emerges a growing number of voices— from diverse geographical locations— pointing GMH’s underlying assumptions in research, policy, and practice in transformative directions avoiding acrimonious debates (Bemme & Kirmayer, 2020; Bemme & Souza, 2014; Kirmayer & Pederson, 2014). Much of the work has been devoted to addressing the boundaries and limits of knowledge production in GMH, as well as embracing political awareness and advocating for an emancipatory agenda. This has highlighted, implicitly or explicitly, the importance of drawing upon critical perspectives to ensure that such efforts are open to diverse worldviews, avoid enacting colonial agendas, and pay special attention to the potential role GMH might play within the hegemonic social order (Applbaum, 2015; Bracken et al., 2016; Ingbly, 2014; Mills, 2014; Mills & Whit, 2017; Summerfield, 2013; White & Shashidhran, 2014).

There are a variety of meanings attached to the notion of “critical” (Kincheloe, McLaren, & Steinberg, 2011). As such, it is imperative to examine how critical approaches have been employed thus far in GMH literature to ensure continuous reflexivity that enables the possibility of a more socially responsive discipline that puts mental health issues to the fore and advances research. It is also pertinent to consider how “critical” approaches have been defined and applied thus far, because critical work is more than just expanding existing theories. Such work is about re-examination of the ontological biases, assumptions, values, and ethics that underpin the field of GMH and re-thinking what may be taken for granted (Hocking & Whiteford, 2012).

In this article, we problematize the standpoints being taken in the published “critical” work in GMH, which considers the existential elements of such claims and raises awareness of underlying assumptions (Bracken et al., 2016; Gómez-Carrill et al., 2020; Sax & Lang,2021; Mills, 2014), will contribute to this overall reassessment of the moral and critical and ethical stances within the field. Citing Foucault (1985), problematization is first and foremost an “endeavour to know how and to what extent it might be possible to think differently, instead of what is already known” (p. 9). From this standpoint, this work invites more scholars to examine knowledge as an object of scrutiny itself, to foster creative and new viewpoints, reflection, and action. Thus, the purpose of this article is to explore and examine how critical perspectives have been taken up in GMH literature thus far to reflect or discuss the possible benefits and challenges of adopting a critical standpoint, while moving forward with an emancipatory or transformative agenda.

## Methodology and Methods

### Methods

Critical Interpretive Synthesis (CIS) is a methodology that aids analysis of complex bodies of literature (Dixon-Woods et al., 2006). Critical interpretive synthesis is an approach to synthesize large amounts of diverse qualitative data that treats literature itself as an object of inquiry, seeking to conduct a fundamental critique rather than critical appraisal (Farrelly & Lester, 2014; Flemming, 2010). This approach was designed by Dixon-Woods et al. (2006) to push beyond compiling and summarizing the literature to engage with the underlying assumptions that shape and inform a given field (Farrelly & Lester, 2014). This approach does not proceed as a linear process with discrete stages of literature searching, sampling, data extraction, critique, and synthesis (Dixon-Woods et al., 2006; Farrelly & Lester, 2014). In contrast, it involves an “iterative, interactive, dynamic and recursive” process (Dixon-Woods et al., 2006, p. 9). It is interpretative rather than aggregative (Bennion et al., 2012) enables critical analysis across diverse forms of literature including qualitative, quantitative, and theoretical papers.

#### Retrieving the Literature and Searching Strategy

We conducted searches across multiple databases, including PubMed, MEDLINE (OVID), PsycINFO, Scopus, and EMBASE, to identify studies published from 2007 to February 2023. We chose 2007 as the starting point, marking the emergence of the Movement for Global Mental Health (MGMH) with influential Lancet papers advocating for global mental health (Patel et al., 2007). Our approach followed the framework for scoping studies outlined by Arksey and O’Malley (2005), which prioritizes an “organic” process to identify relevant material, rather than relying solely on structured relevance searching (Arksey & O’Malley, 2005; Levac, Colquhoun, & O’Brien, 2010). This method was chosen due to the anticipated diversity in the focus and design of relevant articles (Anderson, Allen, Peckham, & Goodwin, 2008).

Two search strategies were used: (1) a combination of abstract, title and key word screening and hand searching, and (2) full-text reading. The first strategy was to review and screen the abstracts, titles, and key words of all 835 articles available online between January 2007 and February 2023. In keeping with Arksey and O’Malley’s (2005) purpose of maintaining a wide approach to generate breath of coverage, no methodological limitations were applied. To be included in this synthesis, the article had to meet one of the following criteria:

a. The article is explicitly defined as critical research or explicitly employs critical perspectives within its theoretical framework, e.g., the authors situate their work within a critical movement or clearly identify their work as a critical study, analysis, etc.
b. The article has an implicit intention to be critical which is not clearly stated, but embedded in the language or purpose of the study using words such as ideology, hegemony, power, or its purpose is to challenge, uncover, reveal, etc.

The search for documents explicitly defined as critical was conducted by screening abstracts, titles and key words using the following electronic search terms: critical, critical theory, critical perspective, critical analysis, hegemony, medical and cultural imperialism, and critical approach. The search for articles that appear to have an implicit critical intention was conducted using hand search, scanning the journal’s content page-by-page, a more organic process of interpretive analysis of the authors’ purpose. Based on the authors’ understanding of critical work, articles were read though to answer the following questions:

- Do the authors intend to question hidden assumptions about global mental health policy, research, and practices or programs?
- Do the author(s) present a critical approach that explains and supports their rationale?
- Do the author(s) aim to challenge any global mental health policy or practice, injustice mechanism or fixed system of thought?
- Are the author(s) using different critical approaches/discourses to critique GMH in the Global South?
- Are the author(s) using different critical approaches/discourses to GMH to provide alternative views to promote mental health care and systems in the Global South?

The initial stage, a combination of abstract, title, and key word screening, as well as hand search, generated eighty articles in total. The second stage was to full text read through the eighty articles searching mainly for the following aspects: the origins and development of critical perspectives in GMH; the focus and content of their critical engagement, how the critical work has been carried out, and how it has pushed the field in certain directions. This broad search provided a range of useful information, highlighting the diverse and powerful contributions made by critical approaches to GMH. At this point, 25 articles were excluded due to their lack of use either implicitly or explicitly of critical approach throughout. Fifty-eight articles were retained in the critical interpretative synthesis (see, ***Table 1 for a chronological list of articles included as a supplementary material)***.

## A Critical Interpretive Synthesis Process

Identifying recurring themes and developing critiques of each of the 54 articles was the first step in the interpretive synthesis and critique process. Each article was critically examined and mapped in relation to its purpose, assumptions, theoretical influence, rationale for the critical approach, and internal, external, and/or broader considerations. Interpretation was iterative and inductive. This involved integrating diverse findings and analyzing each article independently and in relation to others, initial groups of articles were formed. A constant comparison and review of these groupings led to the development of themes illustrating how critical perspectives were incorporated and utilized. Throughout, the original studies’ stated intent was maintained and extended (Dixon-Woods et al., 2006; Heaton et al., 2012). This process allowed the authors to identify sub-themes and draw connections between the individual included articles and the overarching themes. Through this iterative process, authors identified patterns and draw the emerged themes about the utilization and incorporation of critical perspectives in the field of GMH.

## Findings

Through a thorough iterative analysis, we have pinpointed the synthetic constructs of both internal and external critiques carried out within the field of Global Mental Health (GMH). We’ve also discerned the intricate interplay between these critiques and the overarching aspirations and calls for a more critical and emancipatory field of study. The following four key themes have emerged: (1) *Turning the critical lens inward, or the making of the “local” as a source of alterity, resistance, and critique; (2) ‘Turning the critical lens outward”, or a political Turn; (3) Pushing for a broader agenda, and (4) The reflexive turn*.

### 1. “Turning the Critical Lens Inward”: The Making of the “Local” as a Source of ‘Alterity’, Resistance, and Critique

The first eight or nine years within the realm of GMH, spanning from 2008 to 2016/2017, were marked by robust critiques focused on its biological reductionism. These criticisms extended to GMH’s foundational elements, including its conceptual, therapeutic, epistemological, and political underpinnings. Notably, a series of critical articles published during this period, authored by scholars like Summerfield (2008, 2012, 2013), Kirmayer & Pederson (2014), Mills (2014), Bracken et al. (2016), and Mills & White (2017), who keenly challenged the fundamental assumptions and entrenched beliefs that had shaped and defined mental health, illness, and well-being in GMH. This collection of critical work was instrumental in deconstructing the established foundations and pushing the boundaries surrounding the very notion of GMH.

The primary focus of these scholarly endeavors often revolved around questioning prevailing assumptions about mental health across diverse cultures and investigating how specific Western-oriented nosology assumptions and beliefs had come to dominate GMH in both research and practice (Ecks, 2016, 2021; Mills, 2014; Mills & White, 2017). A key argument in this discourse centered on the imperative to move beyond individualistic conceptions of mental health and mental illness, instead advocating for a focus on “situated knowledge generation” (Haraway, 1988). In broader terms, the focus of the critic emphasized the themes of decolonization (Mills, 2014; Summerfield, 2013) and the avoidance of “epistemic injustice” (Fricker, 2003). The concept of “epistemic justice” urged recognition of multiple voices and sources of knowledge, underscoring the significance of local knowledge, which might incorporate various and alternative authorities, methodologies, and criteria for establishing evidence (Kirmayer, 2012). The pursuit of epistemic justice aimed to create an inclusive space that welcomed diverse perspectives, acknowledging that definitions of health and the pathways to mental well-being exhibit variations across different cultural traditions. Consequently, these critical scholars have consistently argued for the adoption of a “pluralistic view of knowledge” when shaping measures for mental health outcomes (Kirmayer & Swartz, 2013).

In line with, the core accusation of GMH is ‘exporting’ a Western model of disorder and treatment to LMICs, neglecting cultural diversity in understanding, and responding to mental suffering, and overly medicalizing distress (see, Mills, 2014; Summerfield, 2013; Bracken et al., 2016; Mills & Fernando, 2014). Further, they took issue with the construction of mental distress as a biomedical pathology, arguing that GMH diverted attention from the underlying social and economic determinants of illness in LMICs (Kirmayer & Pederson, 2014). These critiques also disputed the Mental, Neurological, and Substance use (MNS) framework, which combined mental distress with neurological disorders and substance abuse (as seen in Collins et al., 2011), assuming they were all underpinned by pathological neurobiology (Ingleby, 2014; Fernando, 2014). Concerns were also raised about the influence of the pharmaceutical industry, as pharmaceuticals often represented the primary, and sometimes the sole, approach in GMH intervention until recently (Mills, 2014; Applbaum, 2015; Janis, Jenkins & Kozelka, 2017).

Many scholars expressed concerns about the validity of standardized diagnostic instruments when applied in diverse national contexts, the reliability of epidemiological estimates of global mental disorder prevalence, and the applicability of so-called ‘evidence-based programs’ (Ingleby, 2014; White & Sashidharan, 2014; Whitely, 2014; Ecks, 2016). These scholars argued that the GMH agenda tended to marginalize local communities. They suggested that effective interventions should be tailored to the specificities of local cultures, existing healthcare systems, and the needs of local populations (Kirmayer & Pedersen, 2014). Another line of critique was related to the de-contextualization of suffering. For instance, Mills (2014) contended that translating personal accounts of suffering into context-free psychiatric diagnoses involved abstracting symptoms from their personal and social context, framing problems as brain-based disorders rather than signs of disruption in a person’s “lifeworld” (Kirmayer, 2019) – the experiences, activities, and social networks that give human life meaning (Kirmayer & Pedersen, 2014).

As the debate and global discourse intensified, a dichotomy emerged between the ‘Global-Local’ perspective and the focus on challenging the taken-for-granted assumptions and pushing boundaries within GMH. This re-examination of the fundamental assumptions underlying GMH also broadened the conversation to include a wider range of perspectives, including those often excluded from the traditional narrative.

#### 1.1. The Global-Local Distinction

The dichotomy between forms of support reflecting ‘local’ beliefs and practices specific to certain contexts, and ‘global’ approaches that are standardized and universal, has been a central topic of debate in discussions related to Global Mental Health (GMH) since its inception. Some argue that global mental health initiatives pose a threat to indigenous or local practices (Mills, 2014; Fernando, 2014). Notably, Patel (2014) cautions against idealizing indigenous (local) practices, as they can sometimes involve “inhumane treatments” and practices. However, Mills (2014) also contends that individuals in the Global South “deserve better than being urged to stay in their niche in some great cabinet of ethno-psychiatric curiosities” (p. 134).

Scholars from the Global South, with a particular focus on Southeast Asia, have identified several problematic assumptions deeply embedded in GMH (see, Sax & Lang, 2021; Ecks, 2016, 2021; Das & Rao, 2021). They challenged the global narrative of a mental disorder “epidemic.” Even if statistics suggest a genuine increase in mental suffering worldwide, this doesn’t necessarily mean that therapies and interventions should be unquestioningly imported from the West (Mills, 2014; Mills & Fernando, 2014). Sax and Lang (2021) emphasized that movements like MGMH and organizations such as the WHO appeared to “medicate the symptoms of non-Western “others” to make them more economically productive” (p.14). Echoed by others, they stressed that what is more important to help LMICs populations is addressing the social determinants of mental suffering, which can include issues like inequality, prejudice, and violence (Sax & Lang, 2021; Ecks, 2021; Das & Rao, 2021).

As Bauman (1998) has emphasized, the distinction between ‘local’ and ‘global’ has become increasingly fluid and permeable. New and continually evolving connections are formed between ‘local’ and ‘global’ processes. In this respect, Bemme and D’Souza (2014) highlighted the relevance of anthropologist Anna Tsing’s (2005) concept of ‘friction’ for understanding the interplay between the ‘local’ and ‘global’ within the context of Global Mental Health (GMH). ‘Friction’ encapsulates the idea that the seemingly smooth dissemination of ‘universal’ ideas, concepts, and policies across the world is often impeded or slowed down in specific locations. Yet, it’s important to note that this movement, in the first place, arises from the friction that occurs when these ideas gain traction in a particular setting (Tsing, 2005).

Consequently, the relationship between the global and the local was not viewed as a perpetual hindrance or exclusive promotion but rather a dynamic interplay that defies the zero-sum rivalry often portrayed (Bauman,1998; Bemme & D’Souza, 2014). This dynamic interplay between ‘local’ and ‘global’ finds expression in the hybrid concept known as ‘glocalization’ (Robertson 1994) or ‘glocality’ (Escobar, 2001), which acknowledges the syncretic process that occurs as local and global influences interact. From this perspective, the practice of GMH work shifts away from being a debate about the merits of embracing universal categories versus preserving pre-existing local ones. Instead, it becomes a question of exploring the new possibilities that may emerge from the convergence of these two realms.

#### 1.2. Uncovering Taken-for Granted Assumptions Inherited in GMH Foundations

Numerous authors have undertaken comprehensive analyses aimed at uncovering, identifying, and critically assessing long-standing assumptions that have confined the realm of knowledge production in Global Mental Health (GMH). Through their inquiries, scholars have illuminated how values and normative ideals, stemming from the epistemological foundation of mental health and various facets of the political, socio-cultural, and economic environment within which the discipline evolved, have significantly influenced the generation of knowledge and the presumptions regarding the role of evidence-based medicine (Bemme & D’Souza, 2014).

An early example of a direct critique concerning the epistemological constraints of GMH can be found in Mills’s (2014) incisive examination. Her critique primarily centered on the classification of mental disorders, identifying an overreliance on positivist and empirical approaches in comprehending mental health and illness. This overemphasis, according to Mills, has led to medical imperialism and hegemony. Consequently, Mills advocated for a re-evaluation of the implicit assumptions and ideals underpinning biomedical power and values, particularly those aligned with Western ideologies. Mills’s work (2014, 2015, 2019, 2022, 2023) specifically scrutinized the normative ideals that have guided the discipline since its inception, particularly the promotion of concepts related to “normality,” which, in turn, bolster power dynamics within society. In her book, Mills provocatively raised concerns about the problematic assumptions that underlie GMH policies and practices, which have been inherited from Evidence-Based Medicine (EBM) and tend to solidify ideologies while undermining healing practices and erasing alternatives, especially in the Global South (Mills, 2014; Mills & White, 2017).

In line with this critique, several authors have sought to challenge the foci of the discipline of GMH by critically examining the boundaries established by these ingrained assumptions and beliefs associated with Western ideology, GMH models, and other socio-political influences (see, Bracken et al. 2016; Summerfield, 2008, 2013; Ingleby, 2014; Mills & White 2017; Bemme & Kirmayer, 2020). These authors have called into question the individualistic perspective and the narrow emphasis on categorization regarding mental disorders. Many authors have emphasized the necessity for research that explores mental health and illness within their social and political contexts (Kirmayer & Pederson, 2014; Burgess, et al., 2020).

#### 1.3. Pushing Boundaries by Challenging the foci of GMH

Particularly dominant directions have been to move GMH foci beyond individual experiences of mental health, as a fundamental unity of study and care and position mental illness itself as an object of inquiry; and to conceptualize mental health as a socially and politically situated phenomenon. This work has also demanded a shift in underlying assumptions about the nature and root causes of mental illness that are consistent with critical theoretical perspectives, for example, shifting away from an assumption that human beings are authors of their mental illness towards an assumption that mental illnesses are shaped through and within particular social, political, economic, and other contextual forces. As an example, Fernando (2014) proposed a new strand of scholarship for GMH dedicated to generating knowledge based on the argument that most GMH scholarship is founded on Western notions of personhood, healing, and wellbeing.

Within this trend, articles, Burgus et al (2020), Bracken et al., (2016), Fernando (2014), Mills, (2014), Kirmayer, (2012), Kirmayer and Pederson, (2014) and Lund et al, (2011) articulated key directions for this strand of scholarship commensurate with critical foci, such as: placing mental health in relation to its contextual influences; actively seeking out and describing how cultural, poletical, historical, gender-based, generational, socioeconomic, and other factors restrict access to mental health services. Although drawing on diverse theoretical foundations, this work takes-up a critical intent by raising awareness of the failure to adequately address “mental health and ill-health as situated”, that is, the ways in which mental health and illness is shaped within and contributes to the shaping of contextual factors. For example, Kirmayer, (2012), Kirmayer and Pedersen, (2014) and Kirmayer and Swartz (2013) tied the need to extend the GMH research agenda beyond an individualistic perspective to focus on communities and the structural contexts within which they exist to generate knowledge and raise awareness of how sociopolitical processes, residing outside the control of the individual, predicate and perpetuates emotional wellbeing (Tribe, 2014). The narrow focus of GMH may stem from the ways in which mental health care roles and responsibilities have historically been designated. The concern here is that unresolved sources of social injustice and ‘structural violence’ (Farmer et al. 2006) continue to perpetuate mental health difficulties and limit access to sources of support. In turn, researchers or scholars involved in internal questioning have often to turn the critical lens outwards to political agenda.

### 2. Turning the Critical Lens Outwards: Situating GMH in Relation to Power and Injustice/Political Turn

In the field of GMH, a predominant approach has emerged concerning critical perspectives in the context of political shifts. Within this paradigm, the critique extends beyond a superficial analysis of how GMH policies and interventions are influenced by global or contextual factors. Instead, it delves deep into their positioning within the intricate web of power dynamics that shape relationships. This form of critical research and critique seeks to challenge established beliefs, practices, and norms surrounding the definition of mental health issues on a global scale. It has been instrumental in confronting GMH’s ability to address and rectify persisting inequities and injustices by highlighting the inextricable link between mental health and issues of power and social standing. Through this approach, scholars aim to disrupt existing power structures, whether they are rooted in history, colonial legacies, or economic disparities, and open pathways for alternative perspectives and ways of coexisting in the world.

Furthermore, this research endeavors to reframe mental health as a matter of social justice, acknowledging its profound links with problems like racism, gender inequality, and poverty. It places a strong emphasis on amplifying the voices of marginalized communities and working towards a more inclusive and equitable society. An underlying premise guiding this approach is the recognition that questioning exposes the structures of power and dominance that perpetuate oppressive situations. Within this framework, there is the fundamental assumption that mental health is shaped by specific social, political, economic, and contextual forces.

However, it extends beyond this to assert that GMH itself is inherently a political phenomenon. GMH is viewed as a policy and a practice that enacts social power and contributes to the global social order, potentially perpetuating healthcare disparities LMICs.

#### 2.1. GMH as Means of Governing and Maintaining the Social Order

Another common thread in place that has turned the critical lens outwards is seen in the work of authors such as Mills (2014, 2015, 2019, 2022), Braken etal., (2016), Fernardo (2012, 2014), Summerfield (2008, 2012, 2013), Davar (2014, 2016), and Eckes (2013, 2016, 2021), who challenged scholars to critically analyze the ways GMH may be used as a means to enact social power and govern individuals and collectives in the Global South. In particular, Tribe (2014), in her critical analysis of GMH and cultural awareness within Western psychiatric thought and practice, examines some of the assumptions and political underpinnings that influence GMH.

Many critical scholars, including Tribe (2014), Janis, Jenkins, and Kozelka (2017), Applebaum (2015), and Ecks (2021), asserted that within the discourse of GMH, different cultural constructs, health beliefs, idioms, and local methods for coping with distress are often treated as supplementary layers of meaning, rather than being recognized as the central organizing concepts that they are for many individuals in LMCs. These scholars emphasize that the global dissemination of Western psychiatric concepts, even when well-intentioned, can undermine the rich cultural traditions and heritage of many LMICs, potentially resembling a form of neo-colonialism (Tribe, 2014; Summerfield, 2013; Mills, 2014). In their analysis, they drew upon theories of coloniality, cultural imperialism, and discourse analysis to deconstruct contemporary narratives regarding mental health and illness in GMH. These authors argued that GMH can be used to further entrench existing power structures, which can lead to unequal access to resources and services. They also highlighted the potential for GMH to cause harm to vulnerable populations, particularly those in the Global South. Their aim was to illustrate how the shaping of such discourses within power relations aligned with neoliberal rationality created the possibilities of disease mongering and marketing as there has been relentlessly expansion of defining the boundaries of what mental illness is in the West (Mills & White, 2017; Kirmayer, Lemelson & Cummings, 2015).

In this vein, GMH is a cultural and medical imperialism (Mills, 2014; Summerfield, 2013) that resulted in the medicalization of social problems and the pathologization of normal behavior (Mills & White, 2017), enabling the pharmaceutical industry’s increased power and influence (Applebaum, 2015; Jenkins & Kozelka, 2017). Specifically, Mills (2014) explicitly took up a “critical perspective” to focus on institutional practices and historically shaped “psychiatric illness boundaries” that persist in the West. Similarly, Fernando (2014) and Bracken et al. (2016) have argued that critical lenses are required to investigate how global mental health policies can be construed as global social order, which inevitably perpetuates marginalization, oppression, and injustice. Most importantly, Bracken et al (2016) question how GMH is characterised by an almost “evangelical belief” in biomedical psychiatry benefits (Bracken et al., 2016.p.3) They added that the stark reality transcending everything for many people in the non-Western world, is poverty where one quarter of the global population lives in near destitution, and they asked “what is ‘mental health’ in this broken social world?”( Bracken et al., 2016. P.3.)

All these critiques of Western biomedical models of psychiatry, mental health, and mental illness have questioned the GMH movement as it fails to give sufficient attention to the broader historical and political social contexts. For example, several authors suggest that colonialism and imperialism highlight commonalities between racism, sanism, and ableism, facilitated in part by the proliferation of bio-medical psychiatric practices and legal policies (Ekcs, 2021; Smith, 2014; Heaton, 2013; Mills, 2014; Fernando, 2014; Mills & White, 2017). Therefore, the direct adoption of a biomedical paradigm in GMH and its development of international guidelines for the Global South have been subject to contention. This has resulted in heated debate about the appropriateness of the biomedical model for mental health care in the global South. In this literature, the socially constructed categories, including those of age, gender, race, class and disability have come to the forefront, as mental health scholars and practitioners have increasingly paid attention to how the hegemonic social order, expressed in culture, ideology and social organization, governs mental health care (Joseph, 2015; Kirmayer, Lemelson & Cummings, 2015; Kirmayer, 2012; LeFrancLJois, Beresford, & Russo, 2016).

This shift in theoretical lenses has increased awareness of taken-for-granted exclusionary social practices and recognized the influence of power and the global social order on the shaping of mental health care systems. In that way, this work has pushed the agenda to go deeper than the debates over knowledge generation. It has challenged the GMH advocates and leaders regarding the role of GMH policy, research and practice and what type of mental healthcare GMH is aspiring to promote in LMICs (Kirmayer & Pederson, 2014). This work has encouraged a shift away from the traditional research-based approaches to GMH and highlighted the importance of understanding the cultural, social, and political context of the different countries to create more effective policies and practices. Additionally, it has raised questions about the need for more equitable access to mental health services within and across different countries.

### 3. Pushing for a Broader Agenda: Expanding Beyond Critical Questioning to Transformation

In addition to the critical lenses applied to challenge the fundamental assumptions of the discipline and deconstruct the biomedical model, power dynamics, and social order, scholars have also engaged in critical perspectives to question and reshape the broader role of GMH in addressing societal issues in practice, research, and academia. One such aspect of this work involves scrutinizing the perceived dominance of positivist and post-positivist notions of science within the field, as well as evaluating the discipline’s ethical, political, and moral standpoint, and its capacity to address issues related to power and justice towards engaging with “Transformative Paradigms” in Mental Health (see, Whitaker, 2021).

Through these efforts, scholars aim not only to create room for viewing GMH as a platform that either reproduces and sustains unequal power dynamics or serves as a tool for resistance and societal transformation. A fundamental assumption underlying these endeavors is the need to critique the inherited ideas that positivist science is a value-neutral and wholly objective pursuit solely focused on generating knowledge. Such criticism is seen as necessary to establish space for critical and transformative conceptions of mental health practice. It is further assumed that this space will foster novel approaches to thinking, researching, and taking action concerning mental health disparities and injustices on both local and global scales.

GMH predominantly relies on Evidence-Based Practice (EBP) (Thornicroft & Patel, 2014) as its favored “way of knowing.” EBP advocates for the use of rigorously operationalized empirical methods, primarily Randomized Controlled Trials, meta-analyses, and systematic reviews, to build a research foundation guiding professional practices in medicine and related fields. This approach is designed to safeguard against the undue influence of practitioners’ unverified habits, cognitive biases, or vested interests in specific therapeutic methods.

Nevertheless, there are valid concerns raised by many scholars about whether GMH should wholly embrace EBP, for fear of potentially disregarding the undeniable benefits it has brought to the field of medicine (Ecks, 2016, 2021). As Kirmayer (2012) aptly describes it, there’s a risk of falling into “an epistemological melée in which anything goes” (Kirmayer, 2012. p. 253).

#### 3.1. Standardization and Evidence-Based Medicine

Since its inception, GMH has been the subject of vocal criticism, notably regarding its perceived emphasis on biomedical interventions under the banner of Evidence-Based Practice (EBP). Critics have argued that GMH embodies a medical imperialist approach, primarily expanding markets for psychotropic medication (Summerfield, 2012; Mills, 2014; Mills & White, 2017). In response to such accusations, Patel (2014) has pointed out that a significant portion of GMH research has focused on psychosocial interventions. Patel (2014, p. 786) asserts that withholding the benefits of biomedicine because it was ‘invented somewhere else’. Bemme and D’Souza (2014) have contended that the globalization of intervention methods hasn’t been the primary concern of GMH. Instead, they suggest that GMH’s distinctive feature lies in the dissemination and application of specific epistemologies and research methodologies for evaluating interventions worldwide.

The emergence of Evidence-Based Medicine (EBM) and its hierarchical approach to research evidence has significantly shaped standardized procedures for assessing health interventions. However, Thomas et al. (2007) have cautioned against applying positivist methods from the natural sciences to investigate human behaviors and problems. In line with this critique, EBM has also faced criticism for overlooking the social nature of science and obscuring the subjective aspects of human interactions within the medical context (Goldenberg, 2006).

Nearly a decade ago, Greenhalgh et al. (2014) identified several limitations in the EBM paradigm as it was practiced. These included susceptibility to trial bias, a failure to account for multiple contributions to multi-morbidity, and a tendency to favor ‘algorithmic rules’ over reasoning and judgment. Additionally, some have suggested that ‘gold standard’ EBM methodologies may lack the sophistication required to understand cross-cultural nuances in the comprehension and treatment of emotional distress in diverse contexts (see, Summerfield, 2008; Kirmayer & Pedersen, 2014; Kirmayer & Swartz, 2013).

Kirmayer and Swartz (2013) stressed the importance of GMH embracing a ‘pluralistic view of knowledge’ that can be integrated into empirical paradigms guiding GMH-related research. More recently, the notion of mental health interventions as ‘complex’ interventions that interact with the context to influence outcomes has challenged the gold standard of randomized controlled studies (Moore et al., 2015). Researchers have called for new evaluation methods, including the use of qualitative approaches like ethnography to observe these interactions and unintended effects (Kirmayer & Pedersen, 2014; Kohrt et al., 2016; Roberts et al., 2021). These critiques have led to calls for greater ‘epistemic pluralism,’ which, in turn, is expected to give rise to ‘methodological pluralism’ and “new forms of political recognition and engagement” (Kirmayer, 2012, p. 254). This, in turn, has sparked demands for a more critical and socially responsive discipline (Bemme & Kirmayer, 2020; Bayetti et al., 2023; Bemme et al., 2023; Mills, 2023).

Consequently, the ongoing critiques of GMH have paved the way for transformative and emancipatory GMH policies and practices worldwide. Prominent figures in GMH have demonstrated their willingness to engage with this agenda, as evidenced by their emphasis on identifying the movement with ‘mental health’ rather than solely with ‘biological psychiatry’ and its associated epistemologies (Whitley, 2015), as well as their efforts to establish connections beyond conventional mental health paradigms, particularly with development studies (Patel, 2014).

### 4. Reflexive Turn

The evolution of the GMH field has been closely intertwined with the critiques it has encountered, which have highlighted a recent shift toward “reflexivity” and “mutuality” (Bemme et al., 2023; Bayetti et al., 2023). This shift involves delving into questions about what works, where it works, and for whom it works. Currently, GMH places a strong emphasis on understanding the social determinants of mental health, scrutinizing its own fundamental concepts, acknowledging power dynamics and colonial legacies, and exploring the implications of these issues on funding disparities and epistemic exclusions. In line with these developments, a group of scholars has recently called for “mutuality” (Bemme et al., 2023) as a guiding principle in GMH policy and research, emphasizing equitable knowledge production across epistemic and power differences between the Global North and LMICs (Bemme et al., 2023). They stress the need for mutual learning instead of reinforcing unidirectional knowledge transfers. In broader terms, this call to action promotes reciprocity in GMH policy research and practice (White & Sashidharan, 2014). This shift is particularly significant in highlighting funding disparities and epistemic exclusions, which affect not only researchers in the Global South but also individuals with lived experiences. GMH no longer follows a linear trajectory of success but acknowledges its challenges, embraces lessons learned, and advocates for reflexivity. This renewed relevance and reflexive turn position GMH in a more self-aware and pluralistic epistemic stance, fostering critical engagement from within the field itself.

## Discussion

This article commences by emphasizing the pivotal juncture in the developmental trajectory of GMH. A substantial body of knowledge, grounded in internal critique, has pinpointed the discipline’s key boundaries. Concurrently, critical perspectives have extended GMH’s purview beyond its insular domain, into the socio-political domain. At present, scholars grapple with the imperative of harnessing GMH’s transformative potential and addressing injustices and inequities that underlie mental illness. To contribute to the ongoing discourse aimed at cultivating GMH as a critical and socially responsive discipline, we present an analysis of the implications emerging from a critical interpretive synthesis. These implications are encapsulated in the following constructs: the critical turn inwards, the critical turn outwards, and the broader shift towards transformative approaches and reflective turn. In this context, we situate the GMH as it enters to the critical turn towards transformative /reflexive turn: a moment in its history with a great potential to embrace its praxis-oriented potential fully.

The critical turn inwards is visible within GMH literature that has scrutinized internal limitations, delving into uncovering latent assumptions shaping knowledge production and practices (Mills,2014, Summerfield, 2008, 2013; Kirmayer, 2012). These internal critiques signify a continued scrutiny of the motivations underpinning the GMH movement, particularly the applicability of Western psychiatric diagnostics and treatments in diverse cultural contexts. This scrutiny is prompted by compelling evidence demonstrating cultural variations in mental illness prevalence, symptoms, symptom presentation, and treatment-seeking behavior (Benedict, 1934; Kleinman, 1977, 1980; Kirmayer, 2001), and numerous studies (Ferrari et al., 2013; Kessler & Bromet, 2013; Haroz et al., 2017; Kirmayer et al., 2017; Osborn, Kleinman, & Weisz, 2020; Tekola et al., 2021). Although examples of the critical turn inwards appeared to exist across the time frame reviewed in this paper, it seems to have had its origins in the mid-1990s. In fact, numerous critiques of Western biomedical models of psychiatry, mental health, and mental illness have been advanced since the 1960s (Laing, 1960; Foucault, 1965; Ingleby, 1981, 2014; Double, 2006; Hopton, 2006).

Conversely, the critical turn outwards manifests in works that broaden the scope of GMH critique by refocusing attention on global equality and politics. This expansive perspective enhances our understanding of GMH’s contextual embeddedness and its role in perpetuating social and global inequalities (Bracken et al., 2016; Janis, Jenkins & Kozelka, 2017; Tribe, 2014). This scholarship represents a vital contribution to mental health discourse, recognizing the influence of cultural and sociohistorical factors on mental well-being. Additionally, it creates a platform for addressing socio-political and systemic barriers impacting mental health equity both between and within nations.

Critiques of the GMH movement invite reflection on the type of mental health care envisaged for LMICs. Engaging with these critiques has compelled the field to consider socio-cultural contexts and developmental issues (Patel et al., 2018). The Lancet Commission’s recent reframing of mental health within the framework of sustainable development and the advocacy for a dimensional model mapped along the spectrum of distress – disorder – disability (Patel et al., 2018) are inextricably tied to GMH’s critical turn towards transformative approaches. This demand for ethical, social, and moral engagement propels the discipline towards a more critical and socially responsive stance (Bemme et al., 2023), actively endorsing politically informed transformative strategies (Cosgrove et al., 2019; Hailemariam & Pathare, 2020; Tribe, 2014).

The recent transition towards transformative strategies in GMH has significantly advanced the GMH agenda. However, this evolution is not without its share of debates and tensions among practitioners. It is now imperative to engage in a scholarly discourse concerning the changes and values that the discipline should embrace. This includes considering whether GMH can accommodate a broader diversity of scholarship and advance further towards transformative approaches in mental health (Whitaker et al., 2021) and “fostering reflexive deliberation in practice” (Gómez-Carrill et al., 2020).

Several questions and issues have emerged that demand deliberation among GMH researchers and practitioners as they contemplate adopting transformative approaches and reflexive turn. One pivotal query revolves around the discipline’s capacity to incorporate conceptualizations of mental health rooted in paradigmatic perspectives that conceive science as inherently political, rather than a value-neutral endeavor (Tribe, 2014). Another essential question pertains to whether GMH leaders and researchers, situated within various sociopolitical contexts, are prepared to confront the challenges inherent in embracing critical, transformative approaches. This necessitates questioning institutional systems and structures, strategizing for political engagement, expanding partnerships beyond academia, and being receptive to diverse perspectives challenging conventional beliefs about mental health.

Addressing these questions requires courage, creativity, and determination, but the potential for profound, far-reaching changes in the field of mental health is significant. Recent scholarly dialogues promoting transformative approaches in GMH have already initiated positive change and offer a sense of renewed hope of hope. GMH now places significant emphasis on understanding the social determinants of mental health, re-evaluating its core concepts, recognizing power dynamics and colonial legacies, and considering the implications of these issues on funding disparities and epistemic exclusions. Scholars have called for “mutuality” as a guiding principle in GMH policy and research, emphasizing equitable knowledge production, particularly between the Global North and Global South (Bemme et al., 2023). This emphasis on mutual learning over unidirectional knowledge transfer promotes reciprocity in GMH policy, research, and practice.

In line with this, the recent WHO (2022) “World Mental Health Report: Transforming Mental Health for All” calls for a transformative global mental health approach that aims to inform and promote better mental health for everyone, regardless of their location. This report serves as a critical and well-documented reflection on the progress that has been made and the failures that have been encountered in the field of global mental health. It also serves as an essential guide, providing clear indications of the necessary paths and strategies to ensure the urgent transformations required for a more inclusive and comprehensive mental health system on a global scale.

## Future Considerations

Critical perspectives play a crucial role in ensuring that GMH initiatives are not driven by power dynamics and colonial agendas. Instead, they advocate for respecting diverse worldviews and being mindful of the potential for GMH programs to perpetuate existing social hierarchies.

Despite decades of research, uncertainties persist regarding the nature and causes of mental illness. Controversies also surround the effectiveness of biomedical and psychological treatments. A significant area of critique within GMH revolves around questioning the universality of mental health diagnoses and treatments, as well as their applicability to various local contexts (Das & Rao, 2012; Fernando, 2014). As Hailemariam and Pathare (2020) assert, “it is time to reconsider whether the work in the global mental health domain truly reflects a global perspective” (p. 1012). These limitations have prompted calls for greater “epistemic pluralism,” which, in turn, is expected to lead to “methodological pluralism” and new forms of political recognition and engagement (Kirmayer, 2012, p. 254).

This analysis of critical literature in GMH aims to bring attention to the fundamental assumptions and assertions made in such work. It seeks to explore how critical perspectives and research have been and can continue to be integrated into the field. Recognizing that critical approaches have been present in GMH literature for over 15 years, the proposal for a critical interdisciplinary discipline no longer remains theoretical. Instead, it has evolved into a thriving and stable scholarly movement committed to transcending the traditional role of GMH through reflective engagement in knowledge generation and addressing persistent mental health inequities within, between, and among countries.

This movement endeavors to adopt a more holistic approach to mental health, one that encompasses not only clinical services but also social and economic support, education, and various forms of engagement. Its primary objective is to create a more equitable and sustainable approach to mental health by addressing the root causes of mental health challenges, rather than merely treating their symptoms. As such, GMH researchers, advocates, and practitioners must actively make room within their discipline for the development of an emancipatory and inclusive field of study. These efforts are essential in empowering GMH to become a potent and effective catalyst for positive change.

## Data Availability

All data produced in the present work are contained in the manuscript

